# Effects of COVID-19 in Care Homes - A Mixed Methods Review

**DOI:** 10.1101/2022.04.14.22273903

**Authors:** C Heneghan, M Dietrich, J Brassey, T Jefferson

**Affiliations:** The University of Oxford; Collateral Global; Trip Database Ltd https://www.tripdatabase.com

## Abstract

The report provides an up-to-date review of the global effects of the COVID-19 pandemic in care homes. We used a mixed methods approach to assess care home mortality by country, how the deaths compared with previous periods, and how excess deaths may be explained. We retrieved national datasets for 25 countries on mortality, 17 cohort studies assessing deaths compared to a previous period, and 16 cohort studies reporting interventions or factors associated with excess mortality.

The COVID-19 pandemic disproportionately impacted those living in care homes at the highest risk for severe outcomes. However, the pandemic only highlighted and exacerbated a long-running problem: underfunding, poor structural layout, undertraining, under-skilling, under-equipping, and finally, lack of humanity in dealing with the most vulnerable members of society.

The 17 cohort studies point to excess mortality worsening during the pandemic. Despite involving vast numbers of care homes around the globe, the quality of the evidence is not good. For example, the majority of the studies infer the cause of extra deaths from the observation window (mainly the spring of 2020) rather than through detailed investigations. This is why we do not draw any clear conclusions about the specific causes of death, apart from noting their significantly high numbers. In addition, we did not review all policy actions since 2020 but note there has been a scarcity of studies since then - an indicator that interest in this problem has waned and likely not been addressed.

Analysis of national datasets for 25 countries shows that care home deaths were, on average, 30% of the total COVID-19 deaths (range: 9-64%). The quality of the current evidence base is limited, short term, and lacks standardised methods to prevent robust countrywide comparisons. Residual excess deaths were also observed, with excess mortality being reported for both COVID-19 positive and negative patients.

Several reported interventions or factors suggest the potential to mitigate the risk in care homes substantially. Interventions that could reduce mortality include improving the care home quality, increasing staffing levels, reducing the number of beds in the facility, employing staff confinement strategies with residents, and improving clinical care such as implementing daily examinations. Some care home solutions like US ‘Green House’ homes, which usually have fewer than 12 beds, may provide crucial insights into the care home problem compared with larger homes. Furthermore, care home residents faced barriers accessing emergency treatments during the pandemic waves. Finally, interventions targeting care homes should be subject to smaller trials given large effect sizes in some studies.

Approximately one per cent of the global population resides in care homes, while care home residents account for nearly one-third of deaths attributed to COVID-19 in the 25 countries studied. Reducing this ratio requires analysing current care home infrastructures, funding models, and incentives for providing high-quality care. The scale of the problem in care homes requires robust evaluation and coordinated strategies to improve outcomes for those most vulnerable to COVID-19. Failure to address these systemic problems could mean global care home populations will be similarly affected by future crises and pandemics.

## Background

The COVID-19 pandemic has disproportionately impacted those living in care homes as they are at the highest risk for severe disease and death. [1] Such residents are more at risk of infectious outbreaks as they often share rooms, live in close quarters, and have frequent close contact with staff who move between potentially contagious patients. [2]

Before the pandemic, care homes were struggling to provide high-quality care. [3] Few countries had plans to protect vulnerable people living in long-term care facilities. As the pandemic unfolded, those in care homes faced barriers accessing emergency and elective treatments, and care home improvements were called for. [4]

Numerous media reports highlighted the scale of the problem and the failings. [5]

Furthermore, in February 2021, an international report reported that the share of all COVID-19 deaths in care home residents was 41% (based on data from 22 countries). [6]

An up-to-date review of the global effects of the COVID-19 pandemic is lacking. Not all countries publish data on care home deaths, and international comparisons are problematic due to differences in testing, recording and reporting of deaths, and definitions used. We, therefore, used a mixed methods approach to review mortality in care homes by country, how the deaths compare with previous periods, and how any excess may be explained.

## Methods

We aimed to address three main questions:

1. What were the mortality rates in care homes by country?
2. How does the mortality in care homes compare with previous periods?
3. What explains any excess mortality in care homes?

To answer question 1 (mortality rates by country), we sourced national data on care home deaths and used the most up-to-date data to report the total number of deaths. We also extracted information on the total number of care home cases when reported. We used the same date as the documented care home deaths to estimate the total number of COVID-19 cases using country-specific Worldometer data (https://www.worldometers.info/coronavirus/). If there were discrepancies between the Worldometer and the national data, we used Worldometer data to calculate the care home cases and deaths proportion (and report the national data as a sensitivity analysis). We also recorded the definitions used for ‘care home residents’ and assessed whether descriptions (e.g. ‘skilled nursing care homes’ versus ‘residential care homes’) affect the reported cases or deaths. We also attempted to collate information on the population in care homes to analyse the total proportion of deaths across care homes and report further details on those countries with more than 10,000 care home deaths.

To address questions 2 and 3, we performed a scoping review using a flexible framework for restricted systematic reviews [7]. We searched LitCovid, and the WHO Covid-19 database using the search terms ‘care homes’ OR ‘nursing homes’ AND ‘mortality’. We also performed a bibliography search of included articles.

To answer question 2 (mortality comparisons by year), we included studies that analysed data on deaths compared to a pre-exposure period that permitted estimated excess mortality. We extracted data on the country, region, study type, COVID study dates, length of COVID analysis, population/setting, the sample of care homes, the effect on mortality, causative factors, information from death certificate review, staff sickness, symptoms, and study limitations when reported. Finally, we calculated the percent relative increase in deaths (excess) and the percent of care home-related deaths when reported.

To answer question 3 (excess mortality causes), we included studies that analysed data on interventions and/or factors that might explain excess mortality. We did not include studies that examined patient demographic factors such as age, frailty, ethnicity, symptoms, or rates of COVID that might explain mortality. We excluded case reports and or reviews and cross-sectional studies or surveys. We extracted data on the country, region, the study type, the intervention/exposure, the COVID date, length of study, population/setting, reported effects effect on staff (numbers/sickness, etc.,) impact on residents, and the reported impact on death and any causative factors along with the study limitations. Our review approach is available on the Collateral Global website: What is a Rapid Review? [8]

**Figure 1.**
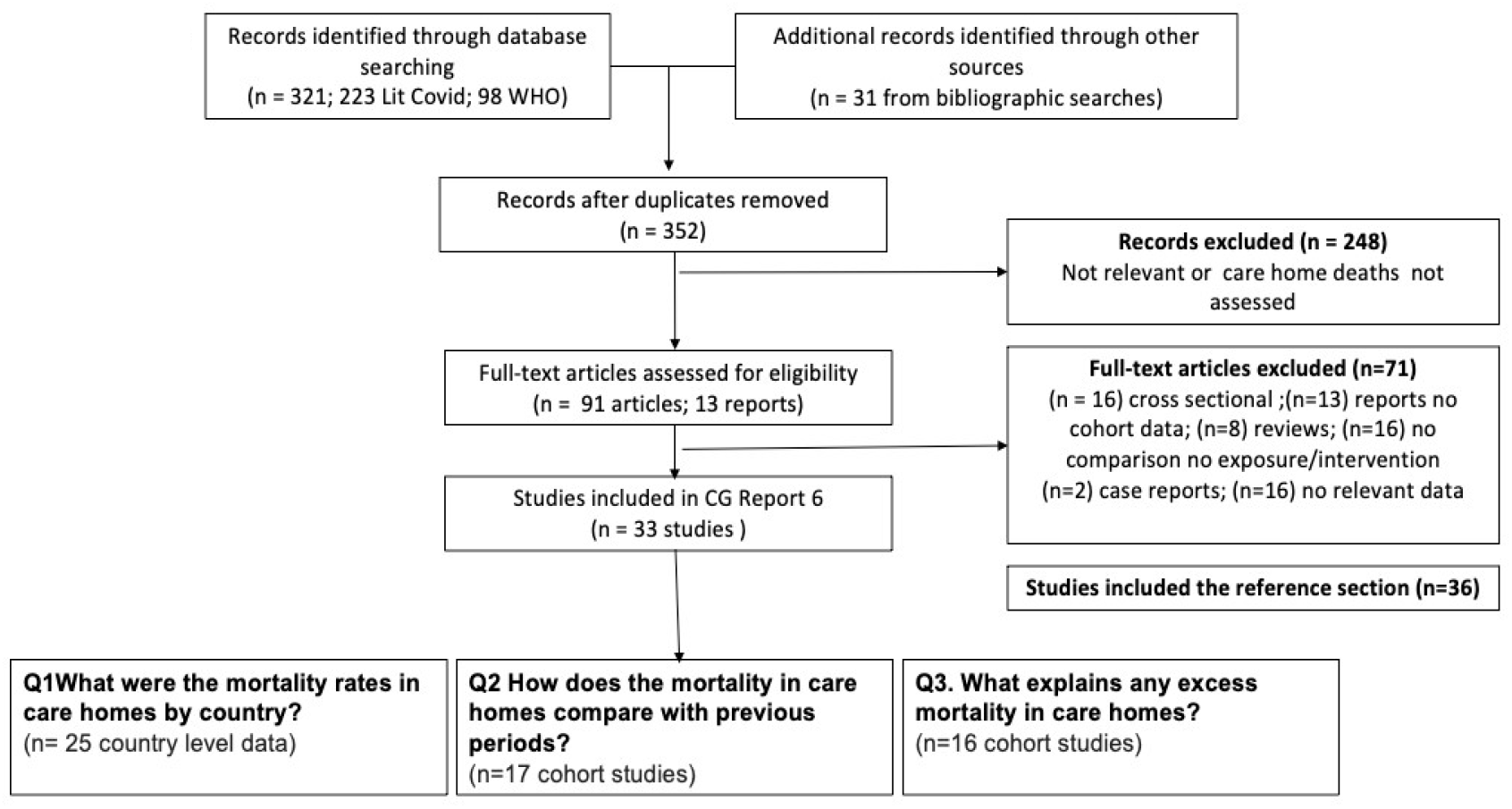
Flowchart for Airborne Transmission.

## Quality

We assessed quality using the Newcastle–Ottawa Scale (NOS) for non-randomised studies, including cohort studies. [9] The scale evaluates 1.) the selection of the cohort, 2.) the comparability of cohorts, and 3.) the outcome. Selection is assessed via the representativeness of the exposed cohort; the selection of the non-exposed cohort, the ascertainment of the exposure, and a demonstration that the outcome of interest was not present at the start of the study. The comparability of the exposed and non-exposed cohorts is assessed based on the study methods. Finally, the outcome is assessed based on objectivity and an adequate length of follow-up.

## Results

### Question 1: What were the mortality rates in care homes by country?

We retrieved data for 25 countries. We also found regional or individual country-level data for two countries, the UK and Canada. End dates for acquiring care home death data ranged from August 2020 (Hungary) to as recent as November 2021 for (Canada) Ontario. We sourced up-to-date data (to September 2021) for Australia, Belgium, Canada (British Columbia), Denmark, Hungary, the UK and the US (See Appendix 1 10.6084/m9.figshare.17104829).

We could not source data from Italy despite widespread problems with staff absenteeism and the need for a public inquest into care homes. [10] Alacevich et al. analysis of care homes and excess deaths during the pandemic in Italy reports official data on COVID-19 deaths in care homes are not available, which prevents accurate estimates of care home deaths. [11] Future updates to care home deaths require a coordinated approach to evaluation. Countries should provide accurate comparative data with regular updates. Given the current variation in reporting and definitions, all comparisons between nations should be treated with caution.

**Australia:** provides data on COVID-19 cases in aged care services/residential care and a weekly report with a data snapshot on the impact of COVID-19 in residential, aged care facilities nationally.

**Belgium:** provides weekly COVID-19 bulletins on COVID-19 monitoring in Belgian nursing and care homes.

**Canada:** provides a COVID-19 weekly report including data on long-term care facilities and retirement residences outbreaks, and British Columbia provides weekly situation reports.

**Denmark:** provides weekly COVID-19 inventories with monitoring data on vulnerable groups, including nursing home residents.

**Hungary:** provides up-to-date information on official COVID-19 measures.

**The UK**: provides Official National Statistics data on COVID-19 on an ongoing basis.

**The US**: reports the Nursing Home COVID-19 Public File, which includes data reported by nursing homes to the CDC’s National Healthcare Safety Network (NHSN) Long Term Care Facility (LTCF) COVID-19 Module.

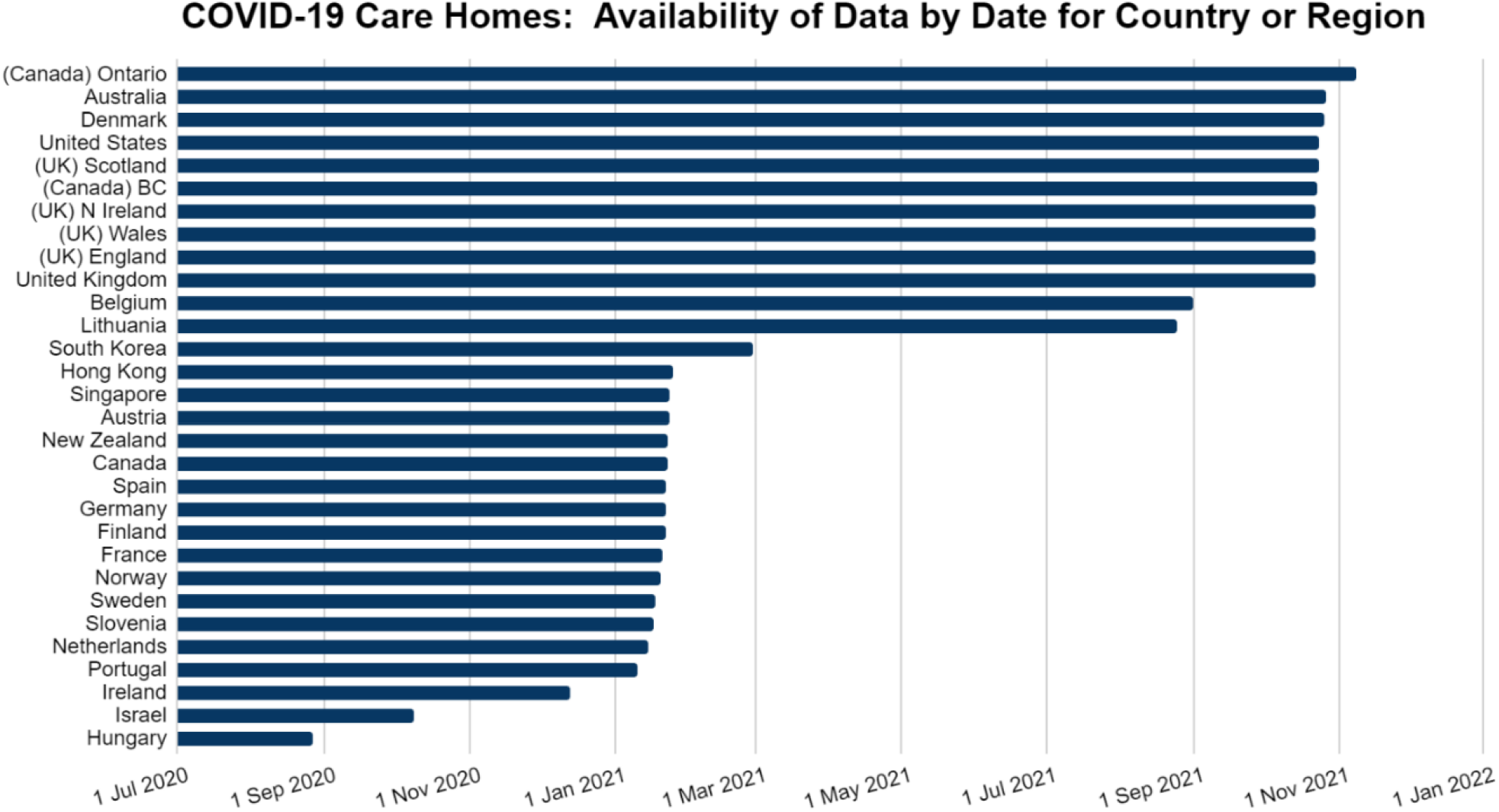

Care home deaths as a proportion of total deaths ranged from 9% in Lithuania to 64% in New Zealand, with a weighted average of 30.0% (median 40%). We found no relationship between care homes deaths as a proportion of total deaths with the total number of population deaths (R^2^ = 0.087) or total cases in the population (R^2^ = 0.091).

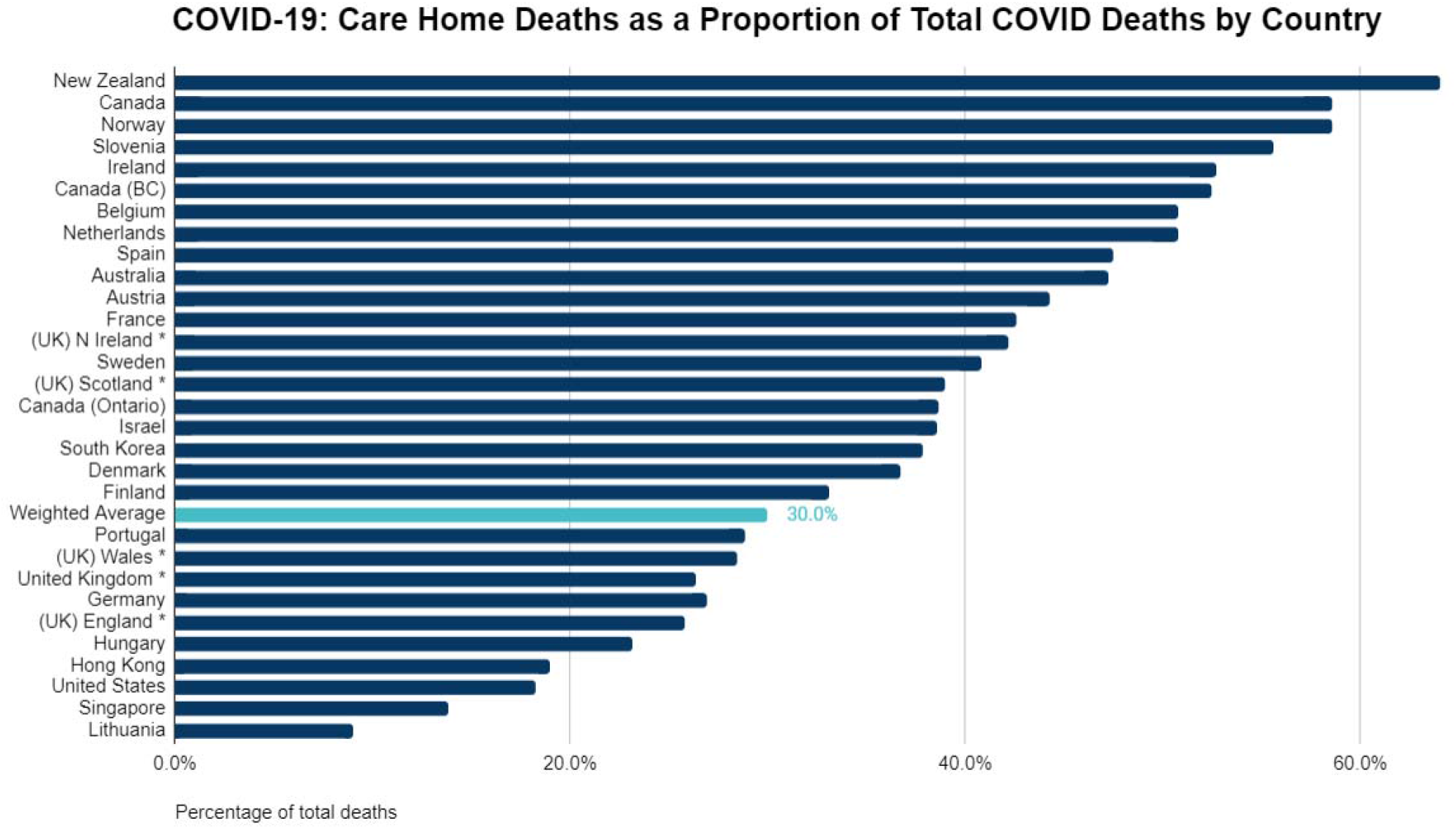

We found considerable variation in the definition of ‘care home’ across countries, which could account for variation in rates of deaths (i.e. description of the setting), and which prevented subgroup analysis by type of residency (e.g., care home, nursing home, long term care facility, or staffing levels, etc.).

Seven countries reported more than 10,000 COVID care homes deaths: Canada, Belgium, France, Germany, Spain, The UK and The US. We summarised these seven countries as follows:

**Belgium**: 12,889 (51%) of 25,380 total COVID-19 attributed deaths occurred in care homes as of 1 September 2021 - Sciensano Link / Link

Belgium has approximately 125,000 individuals aged 65 and over living in care homes. [12] An analysis of data from March to June 2020 in Wallonia (southern Belgium) reported 64% of COVID-19 deaths occurred in care home residents. The COVID-19 mortality rate was 130 times higher inside than outside care homes due to differences in the residents’ age, health frailty, and infection risk. [13] A Médecins Sans Frontières (MSF) report [14] heavily criticised the “alarming living conditions” in Belgian care homes. [15] Nine MSF mobile teams moved from long-term care facilities in early April 2020, identifying problems with a lack of knowledge of basic hygiene rules and treatment protocols and shortages of staff and protective equipment.

**Canada**: 11,114 (64%) of 19,094 total COVID-19 attributed deaths occurred in care homes as of 23 January 2021 - Public Health Agency of Canada. Link

In June 2020, Canada’s national health data agency reported the worst record among wealthy nations for COVID-19-related deaths in long-term care facilities. Approximately 80% of all COVID-19-related deaths in Canada occurred in homes for older people during the first wave. [16] As the pandemic progressed, chronic understaffing in care homes was exacerbated by workers leaving or becoming sick. [17] As a consequence, the army had to be called in. [18]

A study of Ontario residents suggested variations in access to acute care services partly explain the high mortality. Low admission rates in March to April 2020 for care home residents were inconsistent with higher rates seen in subsequent months and stable rates among those living in the community. [19] In June 2020, the Quebec government addressed staff shortages by increasing pay. By July of 2021, 92% of the 9,000 recruited staffers were still working in nursing homes. [20] Since 2021, Canada’s COVID-19 weekly epidemiological update reports a decrease in outbreaks within long-term care facilities. [21]

**France:** 30,395 (43%) of 71,342 total COVID-19 attributed deaths occurred in care homes as of 21 January 2021 - Ministry of Solidarity and Health. Link

As the pandemic progressed, the French government issued several ministerial directives that included a specialist telephone hotline, out-of-hospital mobile geriatric teams, and support for multidisciplinary decision-making. [22] A study of two hospital-dependent French nursing homes compared with a home with no hospital connection suggested high mortality rates might be associated with a lack of medical care management. [23]. Among residents of long-term care in the Southern Ile-de-France region, deaths were mainly due to hypovolemic shock as residents were confined to their rooms for several days without assistance with eating and drinking. As soon as the fifth death was identified, a task force provided medical care including fluids and oxygen therapy, which improved mortality. [24]

**Germany:** 14,066 (27%) of 52,205 total COVID-19 attributed deaths occurred in care homes as of 22 January 2021 - Robert Koch Institute. Link

By July 2020, roughly half of all COVID-19 deaths in Germany were reported to have occurred in care home residents. [25] Germany also suffered from an initial lack of PPE and problems with reduced staff, [26] and there was a reluctance to provide information on care home cases and deaths in some instances. [27]

**Spain**: 26,328 (48%) of 55,411 total COVID-19 attributed deaths occurred in care homes as of 22 January 202 - El Sistema de Monitorización de la Mortalidad Diaria (MoMo) Link / Link

By 27 April 2020, nearly 16,000 deaths (67% of deaths due to COVID-19) were in people who lived in homes or with disabilities. [Cabrero, G] [28] Most were in nursing homes in the autonomous communities of Madrid, Catalonia, and Castile and León. [29] Figures from the Madrid government and the Ministry of Health reported at least 9,470 deaths among nursing home residents in March and April 2020. [30]

In the first wave of the pandemic Spanish armed forces were called up to help nursing homes.

Seniors were found ‘in a state of complete abandonment,’ said the Defence Minister, Margarita Robles. Seventy-one percent of Spanish nursing homes are privately run, and concerns were reported that the system became run down as profit was put before patient needs. [31, 32] In March, nursing homes were placed under the control of public authorities to try to improve the level of care. However, specific policies likely exacerbated the problems. Madrid’s health office instructed staff not to refer residents with COVID-19 infection and severe disability for treatment, and measures such as staffing homes with doctors were rarely implemented. [32]

**The UK**: 37,792 (27%) of 139,811 total COVID-19 attributed deaths occurred in care homes as of 22 October 2021 - Official National Statistics (ONS). Link

In the UK, the Office for National Statistics reports provisional counts of deaths in care homes caused by the coronavirus (COVID-19) by local authorities. The figure shows a sudden sharp rise in deaths in England in week 17 of 2020, a fall and then rise again in the winter of 2020/2021, and then a decline with low numbers of COVID deaths registered up to week 46 (week ending the 19 November). [33]

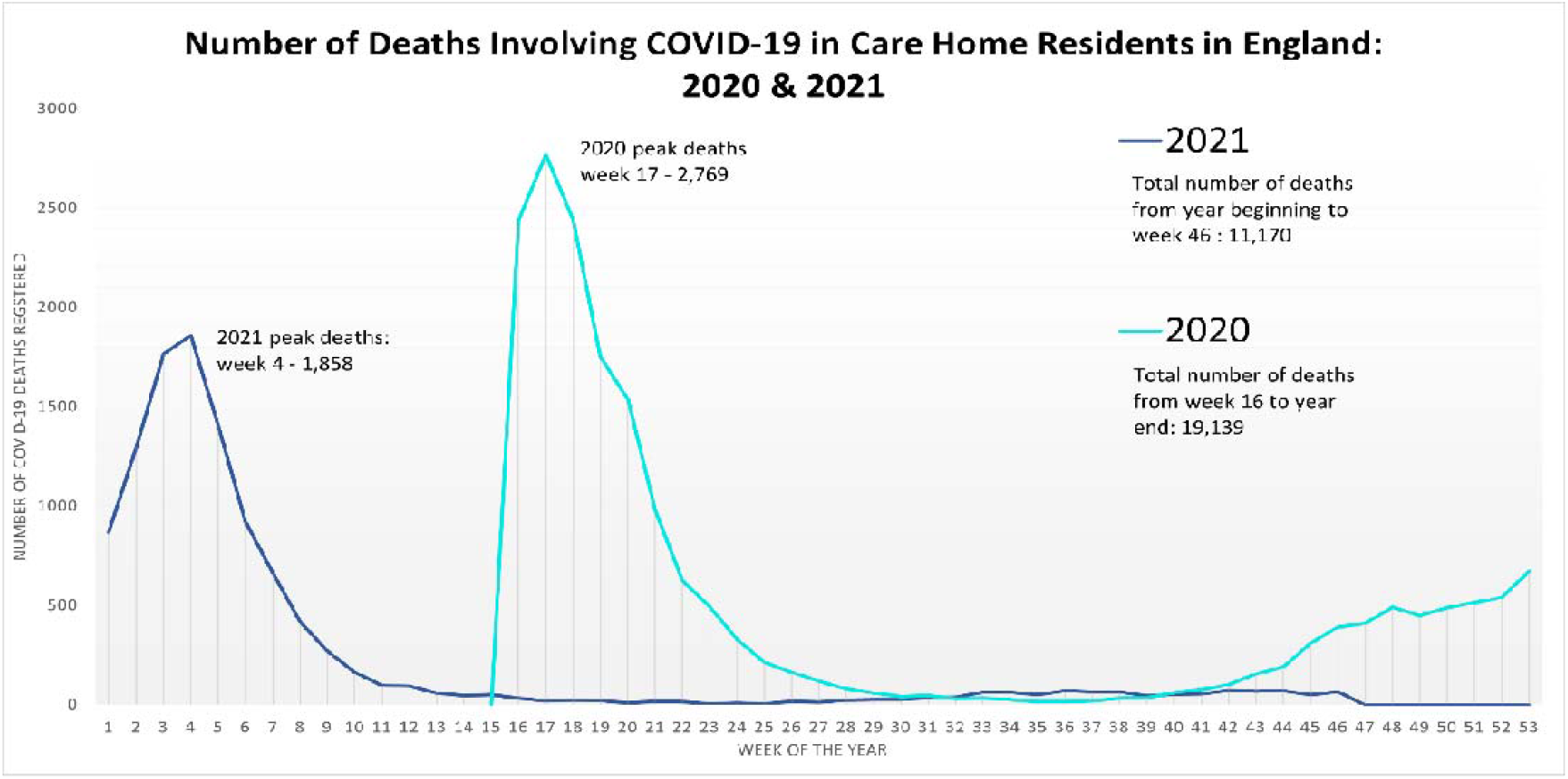

A later ONS report [34] noted a sharp increase in deaths of care home residents involving COVID-19 in the first wave in England and Wales. However, a higher proportion of deaths in care homes was seen in wave two (25.7%) compared to wave one (23.1%), even though excess deaths were higher in wave one (27,079 excess deaths) compared with wave two (1,335 excess deaths).

The ONS suggested the higher number of excess deaths in wave one could be due to delayed access to care services, lack of PPE, or unavailability of vaccines. Mortality displacement seen in wave one and lower care home occupancy levels in wave two also could have contributed.

A joint inquiry by the UK Government’s Science and Technology Committeeland thelHealth and Social Care Committee reported that a ‘lack of priority attached to social care during the initial phase of the pandemic was illustrative of a longstanding failure to afford social care the same attention as the NHS’ and that the ‘rapid discharge of people from hospitals into care homes without adequate testing or rigorous isolation was indicative of the disparity.’ ‘NHS providers were instructed to urgently discharge all medically fit patients as soon as it was clinically safe to do so, and care home residents were not tested on their discharge from hospital.’ [35] The Department of Health and Social Care (DHSC) action plan was published on 15 April. [36] Evidence also suggested that the lack of regular staff testing meant they were more likely to transmit the disease within the care home setting.

**The USA:** 138,985 (18%) of 760,597 COVID-19 attributed deaths occurred in care homes as of 24 October 2021. Centers for Medicare and Medicaid Services (CMS) Link

US data include the Nursing Home COVID-19 Public File that reports to the CDC’s National Healthcare Safety Network (NHSN) Long Term Care Facility (LTCF) COVID-19 Module. The site (https://data.cms.gov/covid-19/covid-19-nursing-home-data) provides data visualisation tools [37] on resident cases and deaths by state.

An analysis by the New York Times in June 2021 reported at least 184,000 COVID-19 deaths among residents and employees of nursing homes and other long-term care facilities. [38] In at least five states, half (or more) of the COVID-19 deaths were linked to care homes, and the case fatality rate (CFR) was reported to be 10%, five times higher than the population CFR.

### National Care Home Data

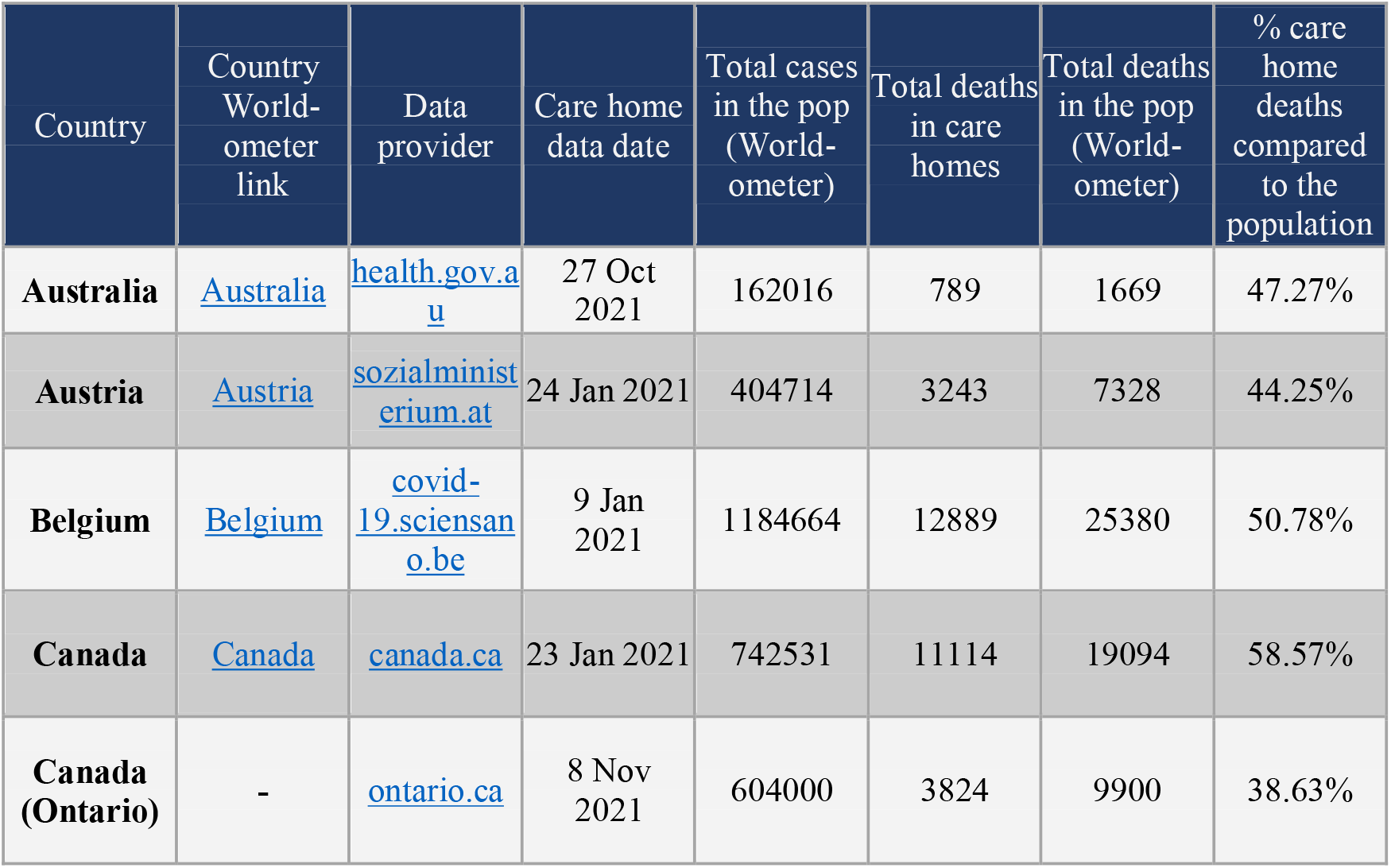
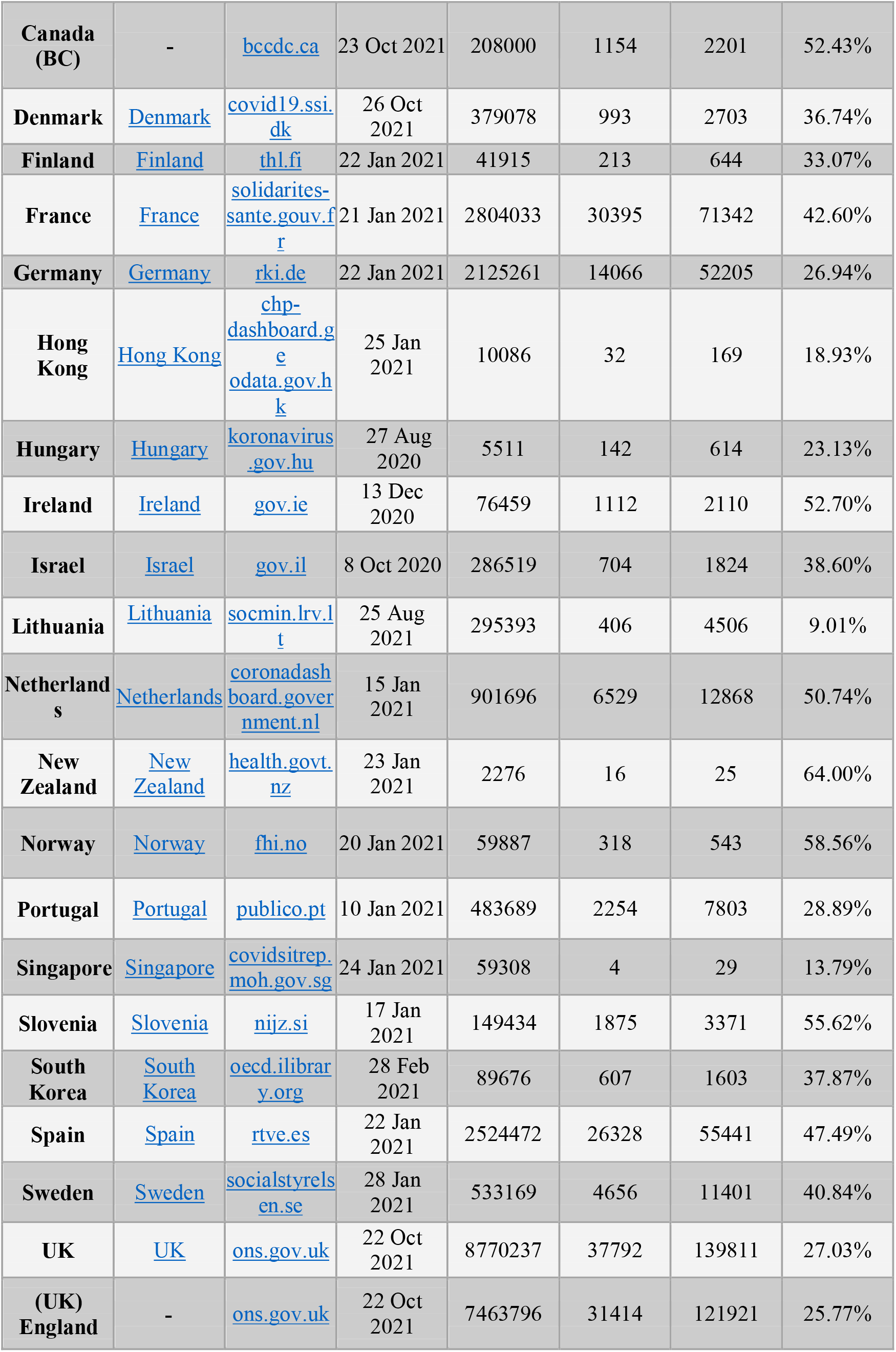
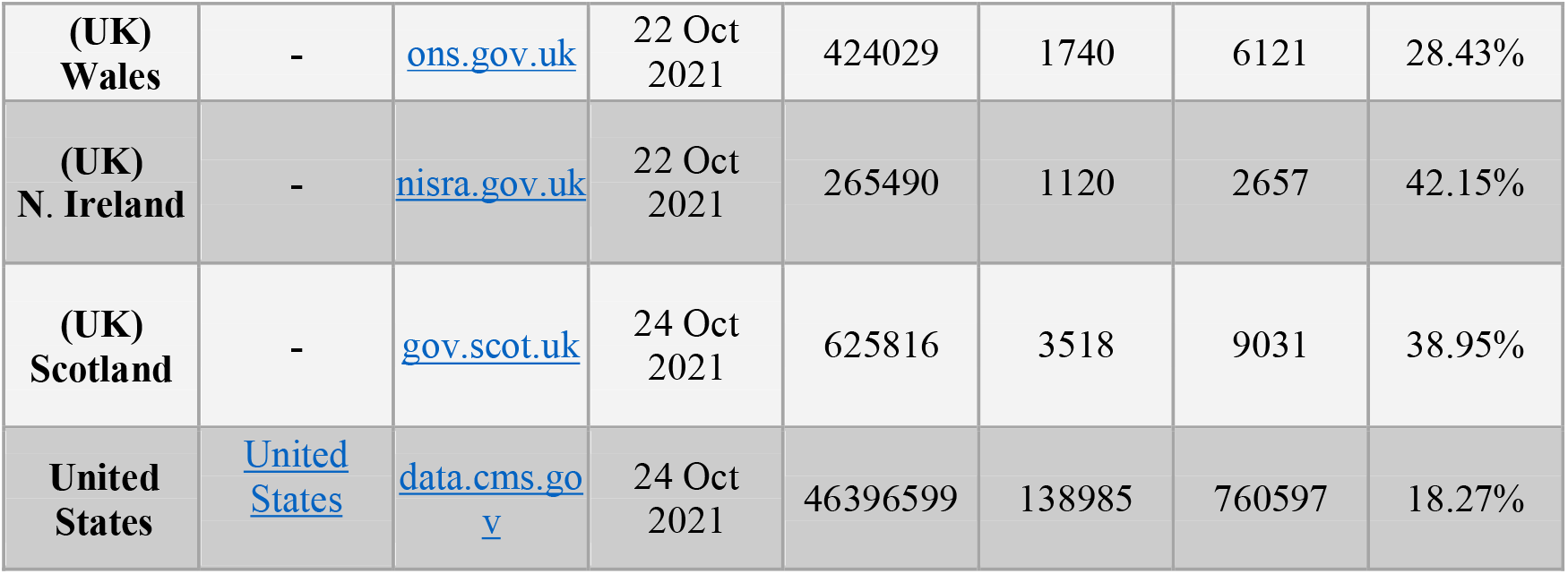

### 2. How does the mortality in care homes compare with previous periods?

We found 17 cohort studies that assessed excess deaths compared to a previous period. Table 1 reports the Excess Deaths Study Characteristics and Table 2 the Care Homes Excess Deaths Study Outcomes (see 10.6084/m9.figshare.17104829)

Three were done in Italy [Alacevich 2021, Ballotari 2021, Cangiano 2020]; two each in England

[Davies 2021, Shoaib 2021], the UK [Graham 2020, O’Donnell 2021] and Canada [Decarie 2021, Sundaram 2021]; and one each in Scotland [Burton 2021], Ireland [Cusack 2020], France

[Canoui-Poitrine 2021], The USA [Greenwald 2021], Wales [Hollinghurst 2020], England & Wales [Wu J 2021] and one in Sweden [Modig 2021]. We also retrieved one multi-country cohort [Signorelli 2020] done in Belgium, Denmark, France, Italy, Spain, Sweden, The UK and The USA.

All 17 studies were observational and therefore prone to bias. The main limitations were the ascertainment of the exposure. Ten studies did not report how the COVID-19 diagnosis was made, and the assessment of the outcome and the follow-up were often inadequate. Studies were short in duration (median four months of follow-up, range: 2-12 months).

All studies report excess mortality during COVID waves 1 and/or wave 2, (see 10.6084/m9.figshare.17104829 for further tabulated results).

**Table 3.** Quality Assessment: Care Home Excess Deaths Studies: Newcastle Ottawa Scale.

| Study ID<br>(Author, Year) | Selection |  |  |  | Comparability of Cohorts |  | Outcome |  |  |
| --- | --- | --- | --- | --- | --- | --- | --- | --- | --- |
|  | Representa-<br>tiveness of<br>the exposed<br>cohort | Selection<br>of the non-exposed<br>cohort | Ascertainment<br>of exposure | Outcome<br>not<br>present at<br>start | Design | Analysis | Assessment<br>of outcome | Follow-up long<br>enough | Adequacy<br>of follow<br>up |
| Alacevich<br>2021 | Yes | Yes | No | Yes | Yes | Yes | Yes | Yes | Yes |
| Ballotari<br>2021 | Yes | Yes | No | Yes | Yes | Yes | Yes | Yes | Yes |
| Burton<br>2021 | Yes | Yes | Yes | Yes | Yes | Yes | Yes | Yes | Yes |
| Cusack<br>2020 | Yes | No | Yes | Yes | Yes | Yes | No | Yes | Yes |
| Cangiano<br>2020 | Yes | Yes | Yes | Yes | Yes | No | Yes | Yes | Yes |
| Canoui-Poirine<br>2021 | Yes | Yes | No | Yes | Yes | Yes | Yes | Yes | No |
| Davies<br>2021 | No | No | No | Yes | Unclear | No | Unclear | Yes | No |
| Decarie<br>2021 | No | Yes | No | No | Yes | Yes | Yes | Yes | No |
| Graham<br>2020 | Yes | Yes | Yes | Yes | Yes | Yes | Yes | Yes | No |
| Greenwald<br>2021 | Yes | No | No | Yes | Yes | Yes | Yes | Yes | No |
| Hollinghurst<br>2020 | Yes | Yes | No | Yes | Yes | Yes | Yes | Yes | No |
| Modig<br>2021 | Unclear | Unclear | No | Yes | Yes | Yes | No | Yes | No |
| O'Donnell<br>2021 | Yes | Yes | No | Yes | Yes | Yes | Yes | Unclear | Unclear |
| Signorelli<br>2020 | Yes | Yes | Unclear | Yes | Yes | Yes | Yes | Yes | Unclear |
| Sundaram<br>2021 | Yes | No | No | Yes | Yes | Yes | Yes | Yes | Yes |
| Shoaib<br>2021 | Yes | Yes | No | Yes | Yes | Yes | Yes | Yes | No |
| Wu J<br>2021 | Yes | Yes | Yes | Yes | Yes | Yes | Yes | Yes | Unclear |

#### Canada

**Sundaram 2021** study in **Ontario** analysed data from March - December 2020 for approximately 14.7 million residents. Mortality rates rose during March 2020 and were higher in all pandemic months through December 2020. However, a substantial reduction in hospitalisations and Emergency Department visits occurred during 2020 compared to previous years (p < 0.001 for each month).

**Decarie** retrospective cohort analysed data on those dying in and outside of nursing homes in **Quebec and British Columbia** from January 1 to June 30. At the pandemic’s peak, excess mortality increased > 150% in nursing home settings.

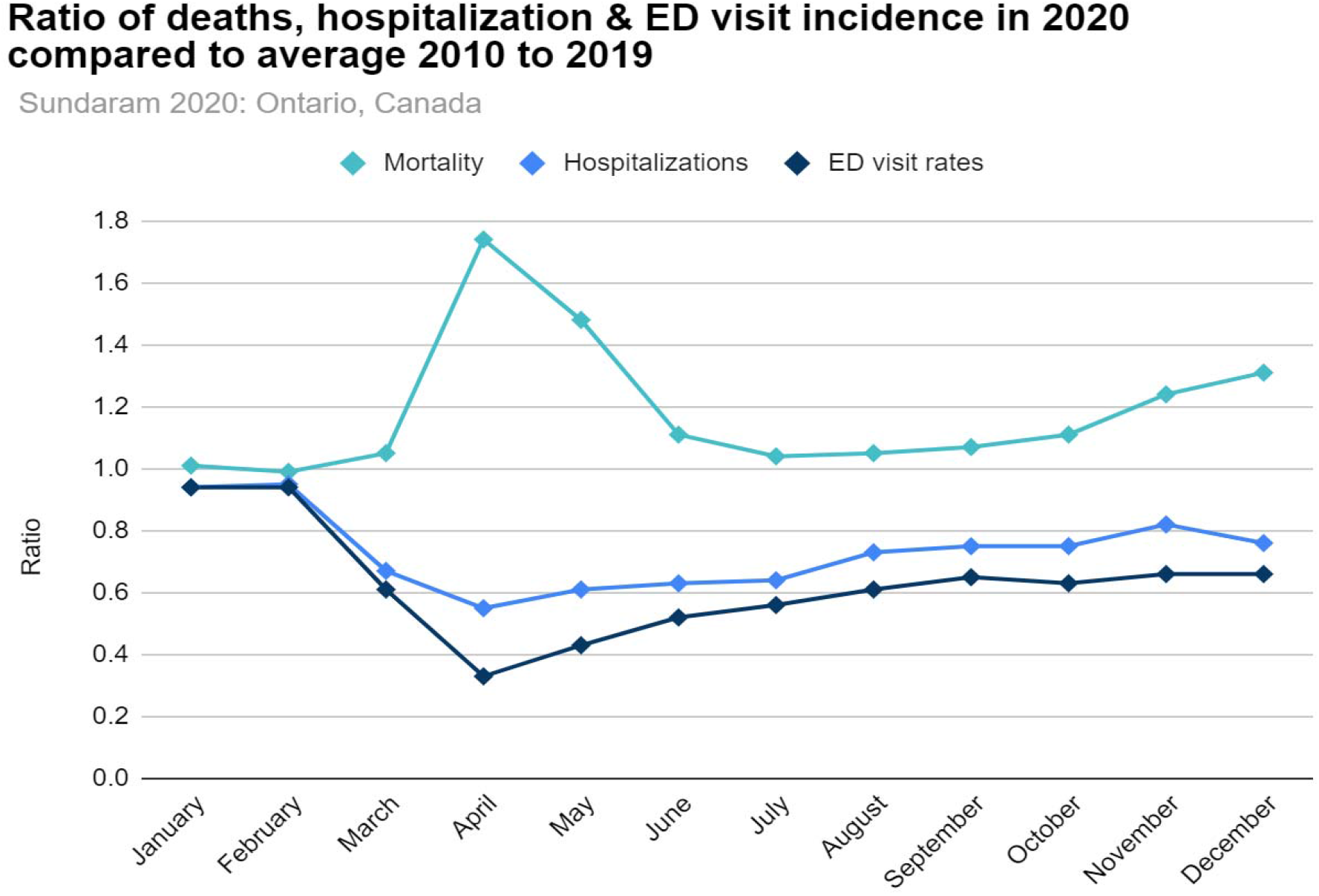

#### France

**Canoui-Poitrine** reported 13,505 excess deaths in a cohort of 494,753 adults in 6,515 **French** nursing homes compared with 2014-19 (mortality increased by 43%, SMR: 1.43). During the first pandemic, the mortality rate among NH residents ranged from 5.3% in Lozère (a rural département with the lowest population density) to 22.2% in the socially deprived Paris suburb, Seine-Saint-Denis.

#### Ireland

**Cusack’s** study using the coroner’s database of death inquiries in the **District of Kildare in Ireland** from March to June 2020 reported an unexplained residual excess of 60 deaths due to natural causes (residual excess 38% compared with 2015-19). Of 139 COVID-19 deaths notified to June 30, 2020, 113 were residents of nursing and residential homes; 25 occurred in the General Hospital (patients admitted directly from the community); and one in the General Community. COVID-19 deaths in nursing and residential homes accounted for 61% of all such deaths in Ireland.

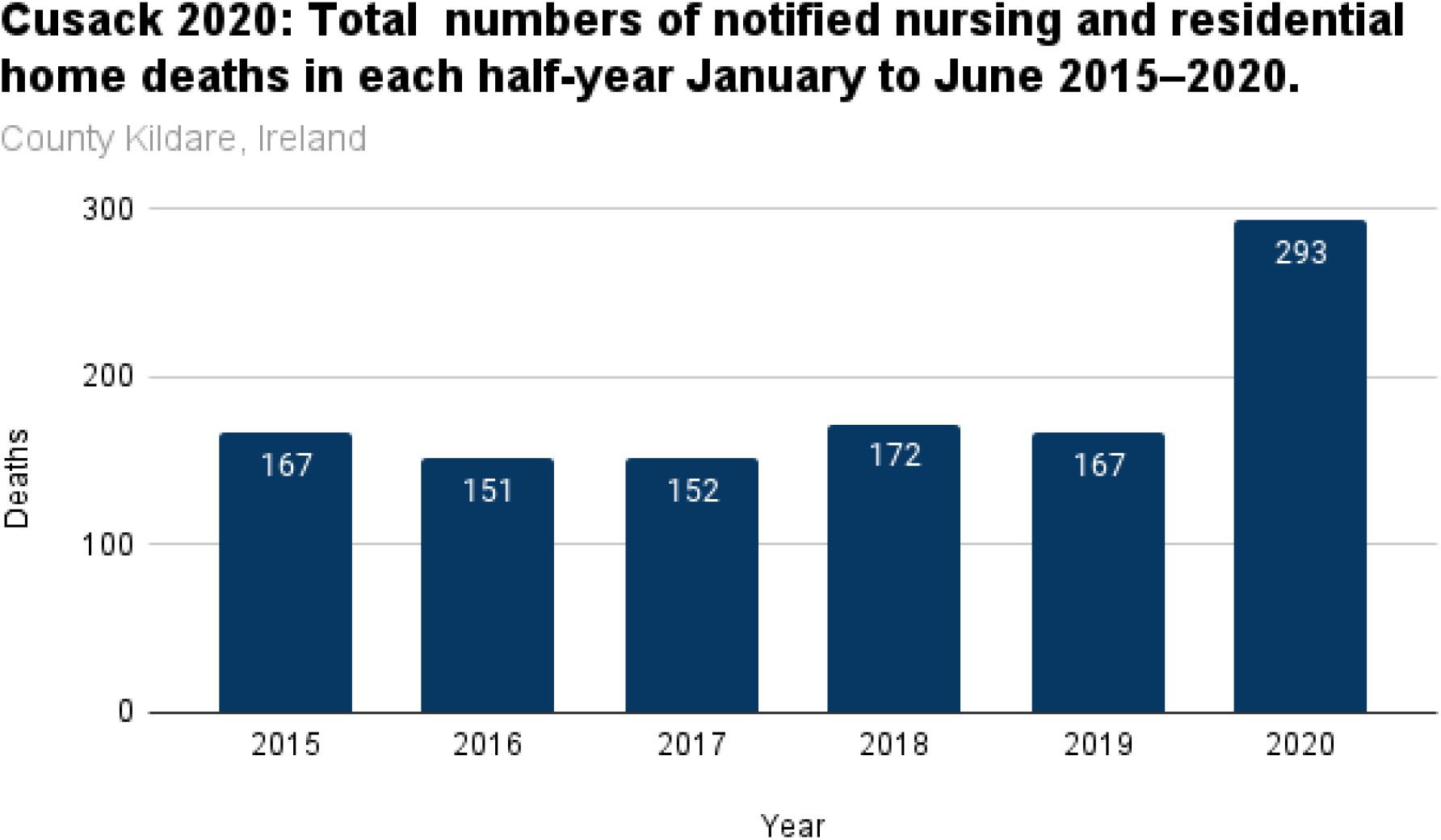

#### Italy

**Cangiano** assessed COVID and non-COVID deaths in an **Italian Nursing home in Milan** over two months and reported mortality was 40% compared to 6.4% in the previous year (63 deaths/157 patients in 2020 vs ten deaths/155 patients in 2019). In addition, an increase was seen in both COVID-19 positive (43%) and negative (24%) patients.

**Ballotari** study in **Northern Italy** in the **Mantua** and **Cremona provinces (Lombardy Region)** reported in the first wave that the nursing care home population excess mortality risk compared to the non-care home population was more than double the 2018 risk, while in a non-nursing care home population, it rose by approximately 60%, and varied by region.

**Alacevich** retrospective analysis between January 1 and March 31 of 2020 in 1,440 municipalities in the **Lombardy Region, Italy**, noted that official data on COVID-19 deaths in care homes for Italy was not available and that the analysis could not differentiate between fatalities that happened inside a care home from the death counts. The results reported that a care home in a region was more relevant than the number of beds in care homes. The data showed that municipalities with care homes in Lombardy registered a higher mortality rate of 6.5%, against a national average of 3.1%.

#### Sweden

**Modig** analysis from April to June 2020 reported excess mortality in care homes, home care & independent living in **Sweden**. Individuals living in care homes experienced the highest excess mortality compared with home care and independent living. During April, the number of excess deaths was higher at every age in the care home group than in the other two groups. However, it was impossible to identify the mechanism behind the excess mortality in the care home groups.

#### The United Kingdom

Burton’s study **in Scotland** reported that 32% of care homes had any cases of COVID-19 up to October 2020. Approximately half of the deaths attributed to COVID-19 were accounted for by the 5% of those over-70s who were care-home residents. Life expectancy fell by almost six months based on care home-specific life expectancy.

**Davies 2021** analysed excess deaths in **England** from March to May 2020 and reported total deaths in care homes were 52,268, 93% above the five-year average (n=27,128). Communities with an increased risk of excess mortality had a high density of care homes and a high proportion of members on income support, living in overcrowded homes and/or non-white ethnicity.

**Hollinghurst 2020** population-based study in **Wales** reported deaths in care homes increased significantly in 2020 compared with the same period of 2016 to the year-end of 2019: adjusted Hazard ratio=1.72 (95% CI, 1.55, 1.90). The study analysed data on Welsh residents (including >12,000 individuals per year in over 500 care homes) from March 23 to June 14, 2020. The analysis adjusted for age, gender, Hospital Frailty Risk Score (HFRS) that led to an increased hazard ratio and the Welsh Index of Multiple Deprivation (WIMD).

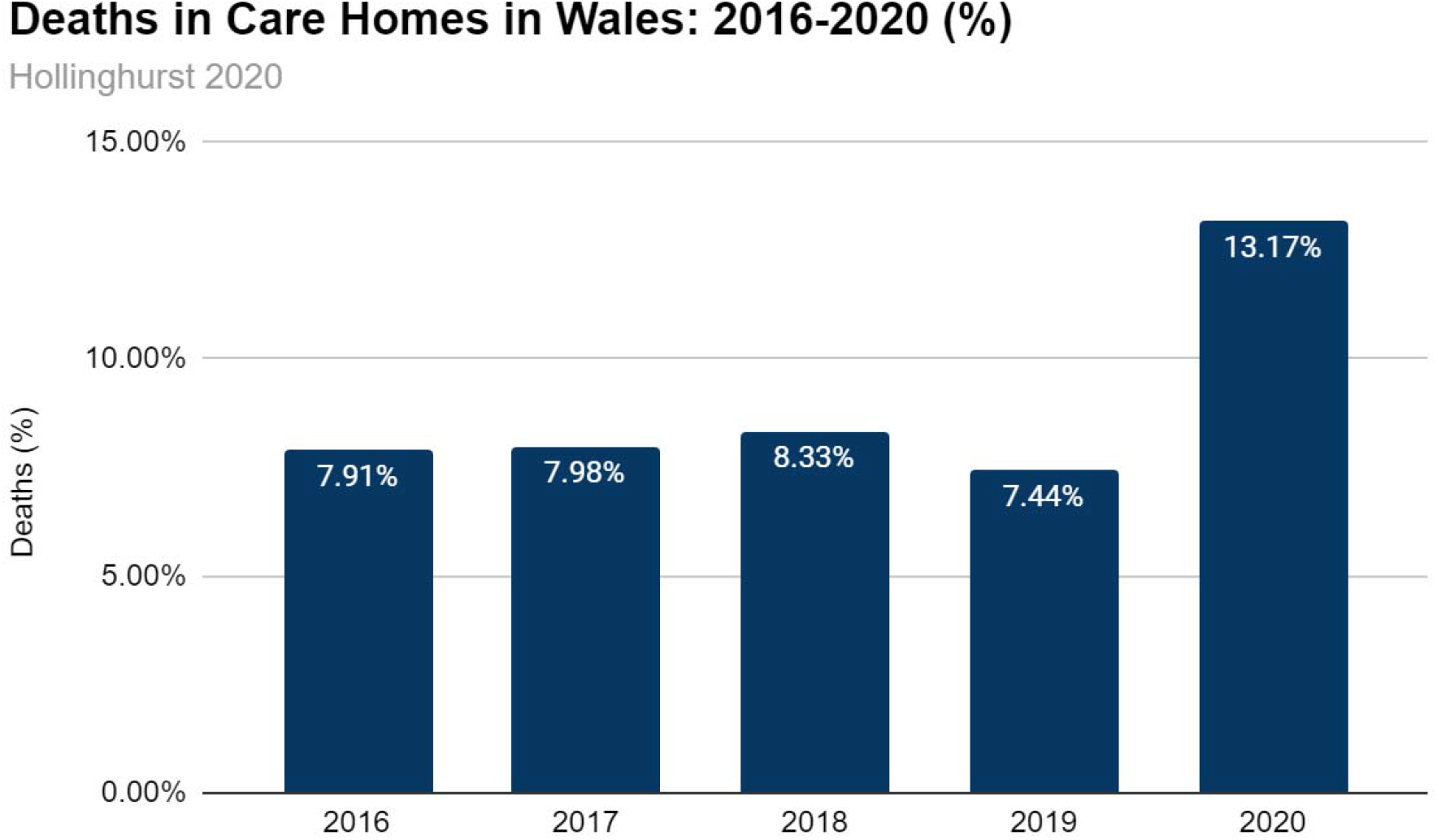

**Wu J** Retrospective Cohort analysed the national death registry of all adult deaths between March 2 & June 30, 2020 in **England & Wales**. Total excess mortality of 57,860 was reported. In care homes or hospices in England and Wales, an excess of 25,611 deaths (44% of the total) was found when compared with historical data between 2014 and 2020: 61% (15,623) of these excess deaths were related to COVID-19.

Of these excess deaths 6,267 were due to dementia, 2,358 to ill-defined conditions in care homes or hospices, of which only 783 and 1,003 were recorded as COVID-19 related. There were 1,495 fewer deaths due to cancer than expected, and 1,211 excess deaths due to cardiac disease in care homes.

Wu et al. noted that care home residents who became unwell during the pandemic may not have been referred to or decided not to go to the hospital for fear of becoming infected.

**In England, Shoaib 2021** reported 36,974 adults with admission and primary diagnosis of heart failure between February 1 and May 31, 2020. There was a 29% decrease in hospital deaths related to heart failure with a concomitant 31% increase in heart failure deaths at home and a 28% increase in heart failure deaths in care homes and hospices (IRR: 1.28, 95% CI: 1.18– 1.40).

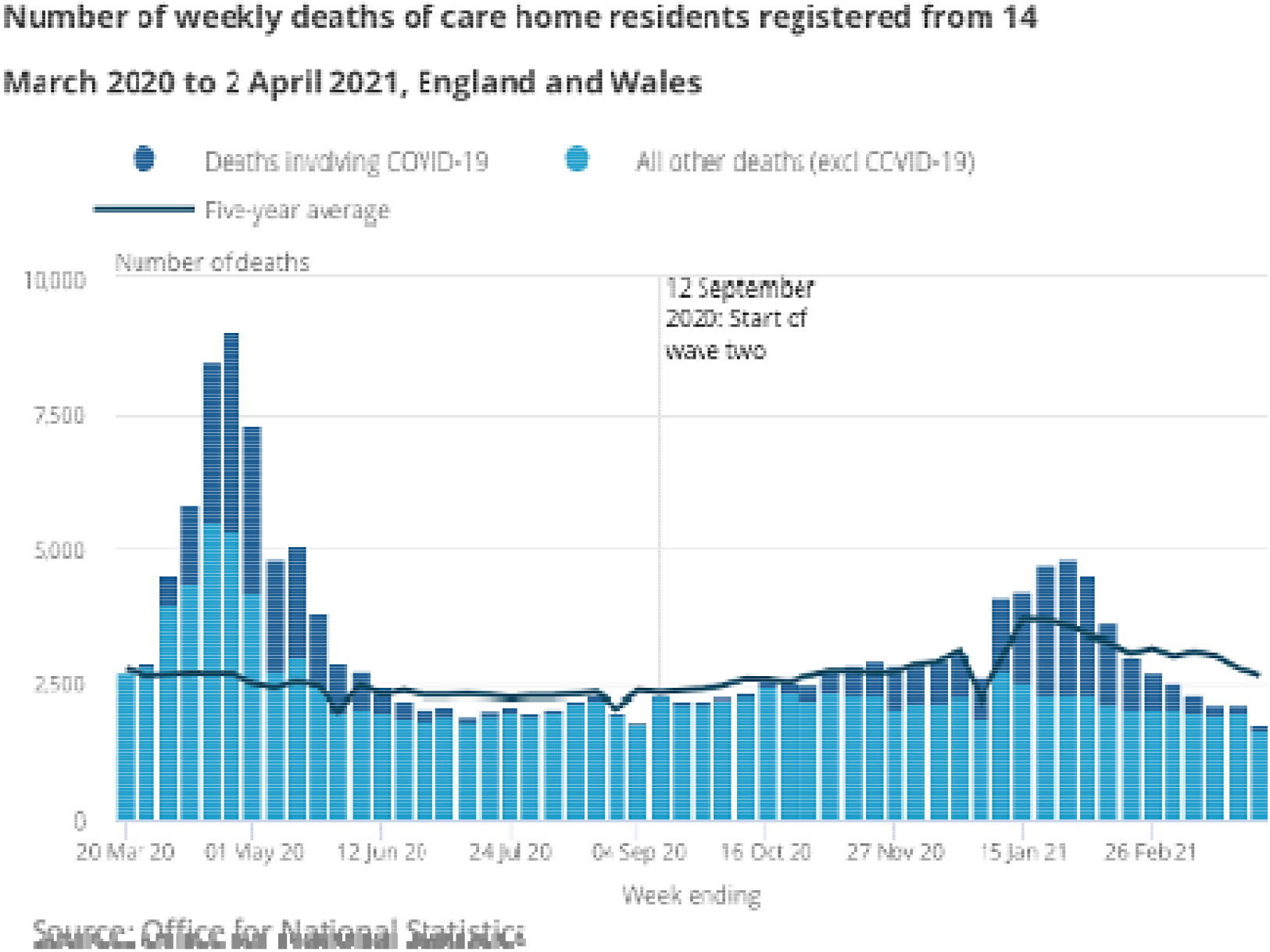

**O’Donnell** analysed the UK’s registered deaths over 12 months (March 2020-21). During the 12 months, 798,643 deaths were registered, of which 147,282 were COVID-19 deaths, and 17,672 were considered additional non-COVID-19 deaths. Deaths in care homes increased above expected levels by 134% during the first wave and 10% in the second wave but fell below expected levels by 3% between waves.

#### USA

**Greenwald** reported on 28,389,098 **US** Medicare and dual-eligible recipients from 29 Feb to 30 Nov 2020 and found that, in long-term care facilities, mortality increased from 20.3% to 24.6% compared with 2017 to 2019.

**Signorelli** reported that the COVID-19 epidemic had seen significant mortality in at least seven of the **metropolitan areas** studied. Different definitions, case ascertainment strategies, and lengths of follow-up will account for variations in reported deaths.

### Signorelli Analysis of Nine Metropolitan Areas from Late Winter-Early Summer 2020

|  | Date | Number of COVID-19 deaths | Proportion of deaths in NHS compared with the region | Non-COVID excess deaths in the population compared with 2019 (%) | Definition of mortality |
| --- | --- | --- | --- | --- | --- |
| New York State | 1 Feb-30 May 2020 | 3,405 confirmed, 2,522 probable | 20% | 8,077/36,644 (21.5%) | People who tested positive and who did not have a positive COVID-19 test, but their death certificate lists as the cause of death "COVID-19" or an equivalent. |
| Bruxelles-Capital | 24 Feb-24 May 2020 | 643 confirmed | 49% | 134/1,455 (9.2%) | Includes both confirmed and probable deaths. |
| Community of Madrid | 1 Jan-24 May 2020 | 1,251 confirmed, 4,728 probable | 69% | 6,325/15,011 (42.1%) | Before 17 April, only people who tested positive were in the hospital. After, independent of the place of death. |
| Catalonia | 1 Jan-24 May 2020 | 4,084 confirmed C19 deaths | 73% | 5,919/11,494 (51.5%) | Before 17 April, only people confirmed and probable cases. After, same as the rest of Spain. |
| Ile-de-France (Paris region) | 11 March-20 May 2020 | 5,621 | 50% | 4,102 /10,952 (37.5%) | Only hospitalised patients who test positive for SARS-CoV- 2 (excluding death occurred in the community or in nursing homes. |
| Greater London | 9 March-16 May 2020 | 803 | 14% | 5,919/11,494 (51.5%) | Only hospitalised patients who test positive for SARS-CoV- 2 (included post-mortem test). |
| Stockholm | 9 March-16 May 2020 | 630 | 32% | 298/2,275 (13.1%) | Patients who test positive for SARS-CoV-2, independently of the cause of death. |
| Hovedstaden (Copenhagen) | 15 March-26 May 2020 | N/A |  | -206/10,741 (-1.9%) | Patients who test positive for SARS-CoV-2, independently of the cause of death. |
| Lombardy (Milan region) | 23 Feb-2 May 2020 | N/A | 50% | 27,334/41,083 (66.5%) | Patients who test positive for SARS-CoV-2, independently of the cause of death. |

### 3. What explains any excess mortality in care homes?

We found 16 cohort studies that assessed interventions or factors that might explain some of the excess mortality reported in sections 1 and 2 of this report. Nine studies were done in the USA [Bui 2020, Collison 2020, Cronin 2020, Das Gupta 2021, Figueroa 2020, Gorges 2020, Harrington 2020, Wang 2021, Zimmerman 2021]; two in France [Belmin 2020, Tarteret 2020] and Canada [Brown 2020, Stall 2020] and one each in Australia [Ibrahim 2021], Germany [Krone 2020] and Spain [Meis-Pinheiro 2021] Table 4 reports the Characteristics of the Studies, Table 5 the outcomes for cases and deaths amongst the studies, and Table 6 the quality assessment (see 10.6084/m9.figshare.17104829).

**Table 6.**
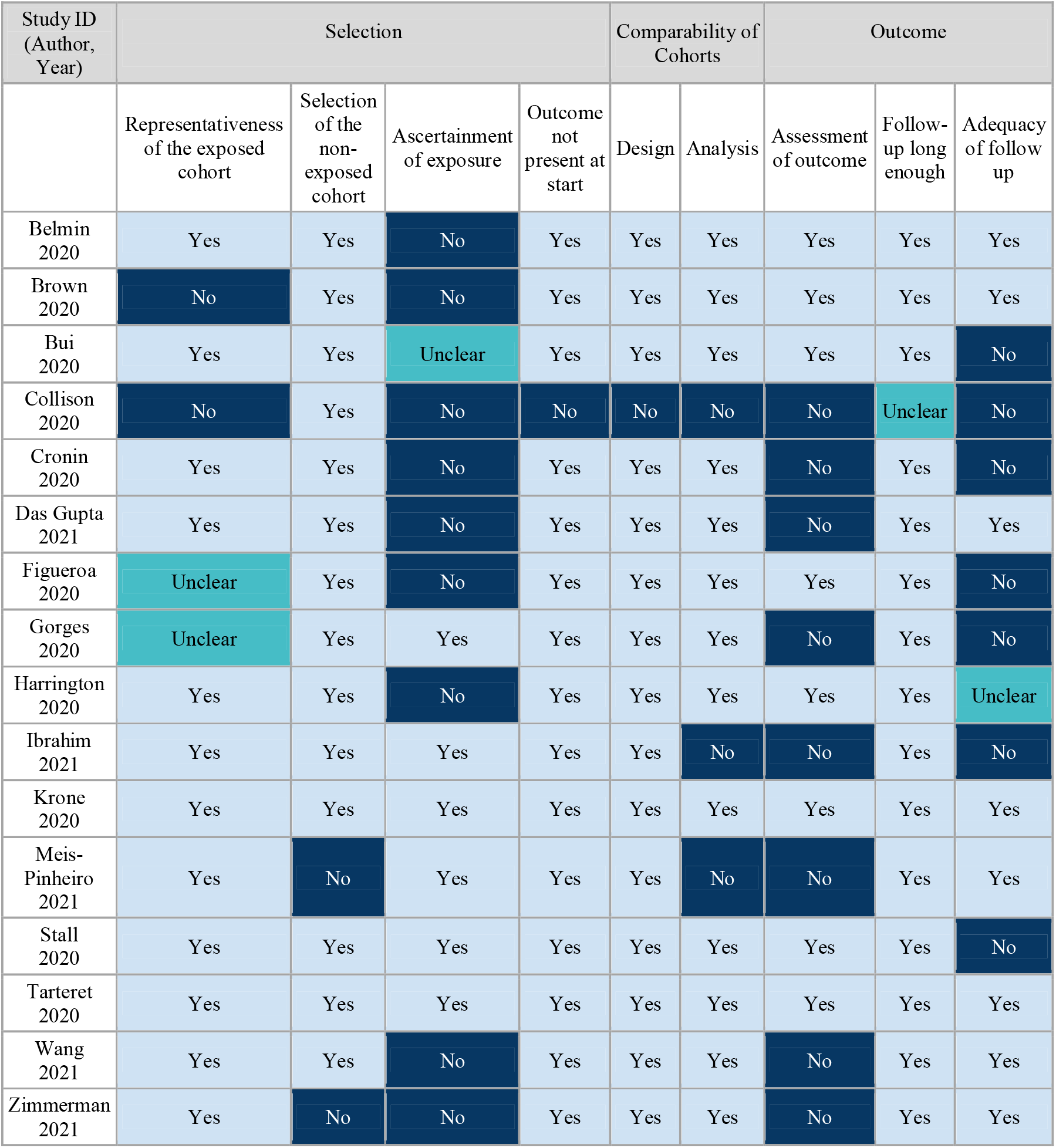
Quality Assessment: Care Home Intervention/Factors Studies: Newcastle Ottawa Scale.

| Study ID<br>(Author,<br>Year) | Selection |  |  |  | Comparability of<br>Cohorts |  | Outcome |  |  |
| --- | --- | --- | --- | --- | --- | --- | --- | --- | --- |
|  | Representativeness<br>of the exposed<br>cohort | Selection<br>of the<br>non-exposed<br>cohort | Ascertainment<br>of exposure | Outcome<br>not<br>present at<br>start | Design | Analysis | Assessment<br>of outcome | Follow-up long<br>enough | Adequacy<br>of follow<br>up |
| Belmin<br>2020 | Yes | Yes | No | Yes | Yes | Yes | Yes | Yes | Yes |
| Brown<br>2020 | No | Yes | No | Yes | Yes | Yes | Yes | Yes | Yes |
| Bui<br>2020 | Yes | Yes | Unclear | Yes | Yes | Yes | Yes | Yes | No |
| Collison<br>2020 | No | Yes | No | No | No | No | No | Unclear | No |
| Cronin<br>2020 | Yes | Yes | No | Yes | Yes | Yes | No | Yes | No |
| Das Gupta<br>2021 | Yes | Yes | No | Yes | Yes | Yes | No | Yes | Yes |
| Figueroa<br>2020 | Unclear | Yes | No | Yes | Yes | Yes | Yes | Yes | No |
| Gorges<br>2020 | Unclear | Yes | Yes | Yes | Yes | Yes | No | Yes | No |
| Harrington<br>2020 | Yes | Yes | No | Yes | Yes | Yes | Yes | Yes | Unclear |
| Ibrahim<br>2021 | Yes | Yes | Yes | Yes | Yes | No | No | Yes | No |
| Krone<br>2020 | Yes | Yes | Yes | Yes | Yes | Yes | Yes | Yes | Yes |
| Meis-Pinheiro<br>2021 | Yes | No | Yes | Yes | Yes | No | No | Yes | Yes |
| Stall<br>2020 | Yes | Yes | Yes | Yes | Yes | Yes | Yes | Yes | No |
| Tarteret<br>2020 | Yes | Yes | Yes | Yes | Yes | Yes | Yes | Yes | Yes |
| Wang<br>2021 | Yes | Yes | No | Yes | Yes | Yes | No | Yes | Yes |
| Zimmerman<br>2021 | Yes | No | No | Yes | Yes | Yes | No | Yes | Yes |

The quality of the evidence was limited. We could not find any randomised trials or controlled studies, limiting the ability to infer causation. Retrospective cohort studies by design are limited in terms of their causative inferences. The study’s main limitation was the ascertainment of exposure, often due to poor reporting.

However, several interventions are worthy of testing in formal trials given large effect sizes and, in some cases, dose effects. Theses interventions include:

| Study ID<br>(Author, Year) | Country | Intervention/Exposure | Decreases in<br>cases | Decrease in<br>deaths |
| --- | --- | --- | --- | --- |
| Belmin 2020 | France | Staff confinement with residents | Yes | Yes |
| Brown 2020 | Canada | Crowding [a] | Yes | Yes |
| Bui 2020 | USA | Quality rating | Yes |  |
| Collison 2020 | USA | Cohorting of positive patients [b] | Yes |  |
| Cronin 2020 | USA | Inspections, staffing, and quality ratings [c] |  | Yes |
| Das Gupta<br>2021 | USA | Quality [d] |  | Yes |
| Figueroa 2020 | USA | Staffing [e] | Yes |  |
| Gorges 2020 | USA | Nursing hours and Aides | Yes | Yes |
| Harrington<br>2020 | USA | Staffing levels [f] | Yes |  |
| Ibrahim 2021 | Australia | Ownership, regulatory noncompliance,<br>location, shared rooms & size | Yes | Yes |
| Krone 2020 | Germany | Separation and screening | Yes |  |
| Meis-Pinheiro<br>2021 | Spain | Density [g] | Yes |  |
| Stall 2020 | Canada | Number of residents/ownership, building [h] | Yes | Yes |
| Tarteret 2020 | France | Resources, hospital dependent,<br>daily examination [i] | Yes | Yes |
| Wang 2021 | USA | Facility characteristics, quality indicators [j] | Yes | Yes |
| Zimmerman<br>2021 | USA | House/Small nursing homes<br>“non-traditional” due to size<br>(10–12 beds) [k] | Yes | Yes |

## Data Availability

All data included in the review is linked to Figshare, an open access repository. See https://doi.org/10.6084/m9.figshare.17104829.

## Footnotes

**[a]** The incidence in high crowding index homes was 9.7% vs 4.5% in low crowding index homes (P□<□.001), and COVID mortality was 2.7% in high crowding vs 1.3% in low crowding homes (P < .001). After adjustment for the regional, nursing home, and resident covariates, the crowding index remained associated with increased infection, RR=1.73 (95% CI, 1.10-2.72) and mortality (RR=1.69 (95% CI, 0.992.87). [Brown 2020]

**[b]** Starting in 2008, the Centers for Medicare and Medicaid (CMS) began providing a “five-star” rating of nursing home quality based on three elements: health inspections, staff-to-resident ratios, and quality metrics such as rates of falls and bedsores, with the first element having the greatest weight in an “overall” rating [Cronin]

**[c]** Compared with 1-star–rated nursing homes, the odds of a COVID-19 outbreak were 87% lower among 2 to 3-star–rated nursing homes, adjusted odds ratio = 0.13 (95% CI = 0.03–0.54) and 94% lower among 4- to 5-star–rated nursing homes, adjOR = 0.06; (0.003–0.39). [Collinson 2020]

**[c]** Nursing Homes with a 5-star inspection rating have a COVID-19 death rate 24% lower than those with a 1-star inspection rating. High-quality NHs were not effective in preventing the spread of COVID-19 among their staff; however, these homes effectively prevented the virus’ spread among their residents – 5-star homes saw roughly 23% fewer cases than 1-star homes. [Cronin]

**[d]** The association diminished over time, showing a reduced role of quality as the pandemic progressed. Suggests that higher-quality nursing homes were better prepared to handle the pandemic in the earlier stages. [Das Gupta]

**[e]** High-performing Nursing homes had a lower number of certified beds. After adjustment, homes with high ratings on nurse staffing were less likely to have more than 30 COVID-19 cases vs facilities with 11 to 30 and vs facilities with ten or fewer cases than low-performing homes, OR = 0.82 (95% CI, 0.70-0.95).

**[f]** Nursing homes with COVID-19 residents were associated with lower CMS nurse staffing and RN ratings than homes without COVID-19 residents. The logistic regression model that considered the effects of health deficiencies, bed size, and ownership reported that the total nurse staffing hours became nonsignificant. [Harrington]

**[g]** The density of the institution showed no correlation with COVID-19 incidence, suggesting homes organised with living units did not have a higher incidence than homes where common spaces were shared. [Meis-Pinheiro 2021]

**[h]** For-profit status was associated with both the extent of an outbreak in an LTC home (adjusted risk ratio [RR] 1.96, 95% CI 1.26–3.05) and the number of resident deaths (adjusted RR 1.78, 95% CI 1.03–3.07) [Stall 2020]

**[i]** Mortality in COVID-19 patients decreased if they had a daily clinical examination, OR: 0.09 (95% CI, 0.03– 0.35, p = 0.01), or vital signs measurement per day, OR: 0.06 (0.01–0.30, p = 0.001). [Tarteret 2020]

**[j]** Compared to NH with >120 beds, those with < 60 beds (ORs = 0.13–0.20); 61–120 beds (ORs = 0.27–0.53), were less likely to have cases. Higher average occupancy increased cases (ORs = 21.24–31.19). Facilities with <60 beds were less likely to report resident COVID-19 deaths. Facilities cited for IPC deficiency >once were more likely to report deaths in all homes. (OR = 1.62, 95% CI [1.11, 2.38], p < .05). [Wang 2021]

**[k]** Rates for all outcomes were significantly lower in Green House/small NHs than in traditional NHs that had <50 beds and ≥50 beds (log-rank test P < .025 for all comparisons). Notably, residents in Green House homes receive significantly more hours per day of care from certified nursing assistants than residents in traditional nursing homes. [Wang 2021]

## Appendix

We plan to update this review based on the emerging evidence.

### Ethics Committee Approval

No approval was necessary.

### Funding

This review received funding from Collateral Global.

### List of Tables available in Figshare

Table 1. Excess Deaths Study Characteristics

Table 2. Care Homes Excess Deaths Study Outcomes

Table 3. Quality Assessment: Care Home Excess Deaths studies: Newcastle Ottawa Scale

Table 4. Care Home intervention/exposure studies characteristics

Table 5. Care Home intervention/exposure studies outcomes

Table 6. Quality Assessment: Care Home Intervention/Exposure studies: Newcastle Ottawa Scale

Figure 1. Flow chart

### Protocol available at Figshare

Appendix 1 National Data

### Competing Interest Statement

TJ’s disclosure is available here: https://restoringtrials.org/competing-interests-tom-jefferson/. CH holds grant funding from the NIHR, the NIHR School of Primary Care Research, the NIHR BRC Oxford and the World Health Organization for a series of Living rapid reviews on the modes of transmission of SARS-CoV-2 reference WHO registration No2020/1077093. He has received financial remuneration from an asbestos case and legal advice on mesh and hormone pregnancy tests cases. He has received expenses and fees for his media work, including periodic payments from the BBC, The Spectator, and other media outlets. He receives expenses for teaching EBM and is also paid for his G.P. work in NHS out of hours and regularly goes into care homes. He has also received income from publishing a series of toolkit books and appraising treatment recommendations in non-NHS settings. He is the Director of CEBM and is an NIHR Senior Investigator. He is co-director of the Global Centre for Healthcare and Urbanization based at Kellogg College at Oxford. He is a scientific advisor to Collateral Global that funds this review. JB is a significant shareholder in the Trip Database search engine (www.tripdatabase.com) and an employee. He has previously received funding from institutions such as WHO, NIHR. Collateral Global funds MD.

